# Estimation of the number of RSV-associated hospitalisations in adults in the European Union

**DOI:** 10.1101/2023.03.09.23287042

**Authors:** Richard Osei-Yeboah, Peter Spreeuwenberg, Marco Del Riccio, Thea K. Fischer, Amanda Cavling, Håkon Boas, Michiel van Boven, Xin Wang, Toni Lehtonen, Mathieu Bangert, Harry Campbell, John Paget, the RESCEU investigators

## Abstract

**Background:** Respiratory syncytial virus (RSV) is a major cause of lower respiratory tract infections in older adults that can result in hospitalisations and death. Estimating RSV-associated hospitalisation is critical for planning RSV-related healthcare needs for the ageing population across Europe.

**Methods:** We gathered national RSV-associated hospitalisation estimates from the REspiratory Syncytial virus Consortium in EUrope (RESCEU) for adults in Denmark, England, Finland, Norway, Netherlands, and Scotland from 2006 to 2017. We extrapolated these estimates to 28 EU countries using nearest-neighbour matching, multiple imputations, and two sets of 10 indicators.

**Results:** On average, 158 229 (95%CI: 140 865-175 592) RSV-associated hospitalisations occur annually among adults in the EU (above 18 years); 92% of these hospitalisations occur in adults over 65 years. Among 75-84 years old, the annual average is estimated at 74 519 (95%CI: 69 923-79 115) at a rate of 2.24 (95%CI: 2.10-2.38) per 1000 adults. Among adults aged ≥85 years, the annual average is estimated at 37 904 (95%CI: 32 444-43 363) at a rate of 2.99 (95%CI: 2.56-3.42).

**Conclusion:** Our estimates of RSV-associated hospitalisations in older adults are the first analysis integrating available data to provide estimates of the disease burden in this population across the EU. Importantly, for a condition which was considered in the past to be primarily a disease of young children, the average annual hospitalisation estimate in adults was lower but of a similar magnitude to the estimate in young children aged 0-4 years: 158 229 (95%CI: 140 865–175 592) versus 245 244 (95%CI: 224 688 –265 799).

## Introduction

Respiratory syncytial virus (RSV) is a major cause of acute respiratory infections (ARI) in both infants and older adults. In adult populations with RSV infections, lower respiratory tract infection is common and can result in respiratory failure or death [1, 2]. RSV is a common cause of hospitalisation for older adults especially in the winter months and the commonly associated diagnoses include pneumonia and exacerbations of chronic conditions such as chronic obstructive pulmonary disease (COPD), congestive heart failure (CHF), and asthma [3-7]. Severe respiratory illnesses resulting from RSV infections with complications that could be compared to those caused by seasonal influenza, often among influenza-immunised populations, have been reported in hospitalised adults [2]. The mean length of stay among RSV-associated hospitalisations as compared to influenza is longer among adults (6.0 days vs 3.6 days) [8], and an even longer median length of stay of 9 days (6-25 days) has been reported in adults hospitalised with RSV [9].

A high burden of RSV hospitalisations among adults, notably older adults and persons with chronic conditions including transplantation, COPD, and CHF has been reported [10, 11]. Previous studies have reported varying estimates of the incidence of RSV infections in hospitalised adults [5, 11, 12]. Estimating the incidence of RSV infection in hospitalised adults remains challenging as there are no dedicated RSV surveillance systems, RSV is not routinely screened for in patients with an ARI and only the more severe cases are commonly diagnosed [13]. RSV-associated mortality among older adults is reportedly substantial and comparable to influenza [14] with the majority of deaths occurring among persons aged ≥60 years [8]. Global estimates indicate that the RSV-associated hospital admission rate and in-hospital case fatality rate (hCFR) are higher among persons aged ≥65 years compared to those aged 50-64 years [12]. Older adults may be more susceptible to viral and bacterial diseases and complications including RSV-associated hospitalisations partly due to immunosenescence [15] and the number of comorbidities that exist in this population [16].

It is recognised that RSV infections are common in older adults in high-income countries [7, 17]. To effectively plan healthcare resource utilisation and adequately manage the RSV-related healthcare needs of the older adult population across Europe, including prioritising preventive care, it is important to understand the RSV disease burden in this population. In this study, we aimed to estimate the numbers and incidence rates of RSV-associated hospital admissions in 28 European Union (EU) countries including the UK and Norway by using data from a previously published REspiratory Syncytial virus Consortium in EUrope (RESCEU) study [18] and a literature review.

## Methods

We used a two-stage approach to estimate RSV-associated hospitalisations in adults across the countries in the EU.

### Stage 1 input data

In Stage 1, we gathered and used modelling inputs from the previously published RESCEU project estimating nationally RSV-associated average annual hospitalisations and hospitalisation rates for adults in Denmark, England, Finland, Norway, Netherlands, and Scotland from 2006 to 2017 [18]. These studies used time-series regression methods to estimate the RSV-associated numbers and rates of hospitalisation.

#### RSV hospital admission definition in RESCEU studies

In Stage 1, the previous RESCEU studies identified all respiratory hospital admissions using ICD-10 codes at any point during an admission. The studies extracted all hospital admissions with any mention of respiratory tract infection (RTI), RTI admissions with any mention of pathogen-specific diagnosis code (pathogen-coded admissions) and RTI admissions with an RSV diagnosis code (RSV-coded admissions) [18].

#### Scoping literature review

In addition, to complement RESCEU data, we conducted a scoping literature review in November 2021 to identify estimates that used the same methodology in the RESCEU studies. We searched Medline and Embase electronic databases using pre-defined terms to identify original articles on RSV-associated hospitalisations (Supplementary File 1). The search broadly focused on national estimates published in European countries between 2000 and 2021. Our search did not apply any language restrictions. Two independent reviewers (ROY and MDR) conducted title and abstract screening, full-text screening and data extraction. We followed the Preferred Reporting Items for Systematic Reviews and Meta-Analyses (PRISMA) guidelines [19]. The diagnosis codes used to identify respiratory diagnosis groups of the data sources that provided Stage 1 estimates are presented in Table 1.

**Table 1.** Diagnosis codes used to identify respiratory diagnosis groups of the data sources that provided Stage 1 estimates

| Author, year | Country | Period of observation | Age groups | Outcome coding |
| --- | --- | --- | --- | --- |
| Johannesen et al., 2022 | Denmark | 2010-2017 | 0-2m, 3-5m, 6-11m, 1-2y, 3-4y, 5-17y,18-64y, 65-74y, 75-84y, 85+y | ICD-10: Acute upper respiratory tract infection (URTI) (J00, J02-06);<br>Pneumonia & influenza (J09-18);<br>Bronchiolitis and bronchitis (J20-21, J40);<br>Unspecified LRTI (J22) |
| Johannesen et al., 2022 | England | 2007-2017 |  |  |
| Johannesen et al., 2022 | Finland | 2006-2016 |  |  |
| Johannesen et al., 2022 | Netherlands | 2013-2017 |  |  |
| Johannesen et al., 2022 | Norway | 2008-2017 |  |  |
| Johannesen et al., 2022 | Scotland | 2010-2016 |  |  |

#### Stage 2 statistical modelling

In Stage 2, we extracted the data from the six national RESCEU estimates to create the input data. We extrapolated the input data to the EU using two different modelling approaches the nearest-neighbour matching and multiple imputations, and two sets of 10 indicators (Supplementary File 2) [20]. This resulted in four distributions of plausible values for each country. Next, a hierarchical linear model was used to estimate a rate and confidence interval for each country. These approaches generated four sets of estimates, with confidence intervals and the averages of these estimates and the confidence intervals were reported. We estimated the average number of RSV-associated hospitalisations and the annual hospitalisation rates (per 1 000 adult population) by age group in the EU including the UK and Norway. The outputs of the two modelling approaches for the EU are presented in Supplementary Files 3-10.

## Results

### Scoping literature review

Of 1 392 citations assessed, five were identified to be eligible for inclusion in the analysis. Two of the eligible studies provided estimates only for infants and data from these studies were not included in this analysis. We found three eligible studies providing new estimates for England [21-23], but because there were recent RESCEU estimates for England, the new estimates found from the literature were not included as input data for Stage 2. The search records of RSV-associated hospitalisation estimates are outlined in a PRISMA flowchart (Figure 1).

**Figure 1:**
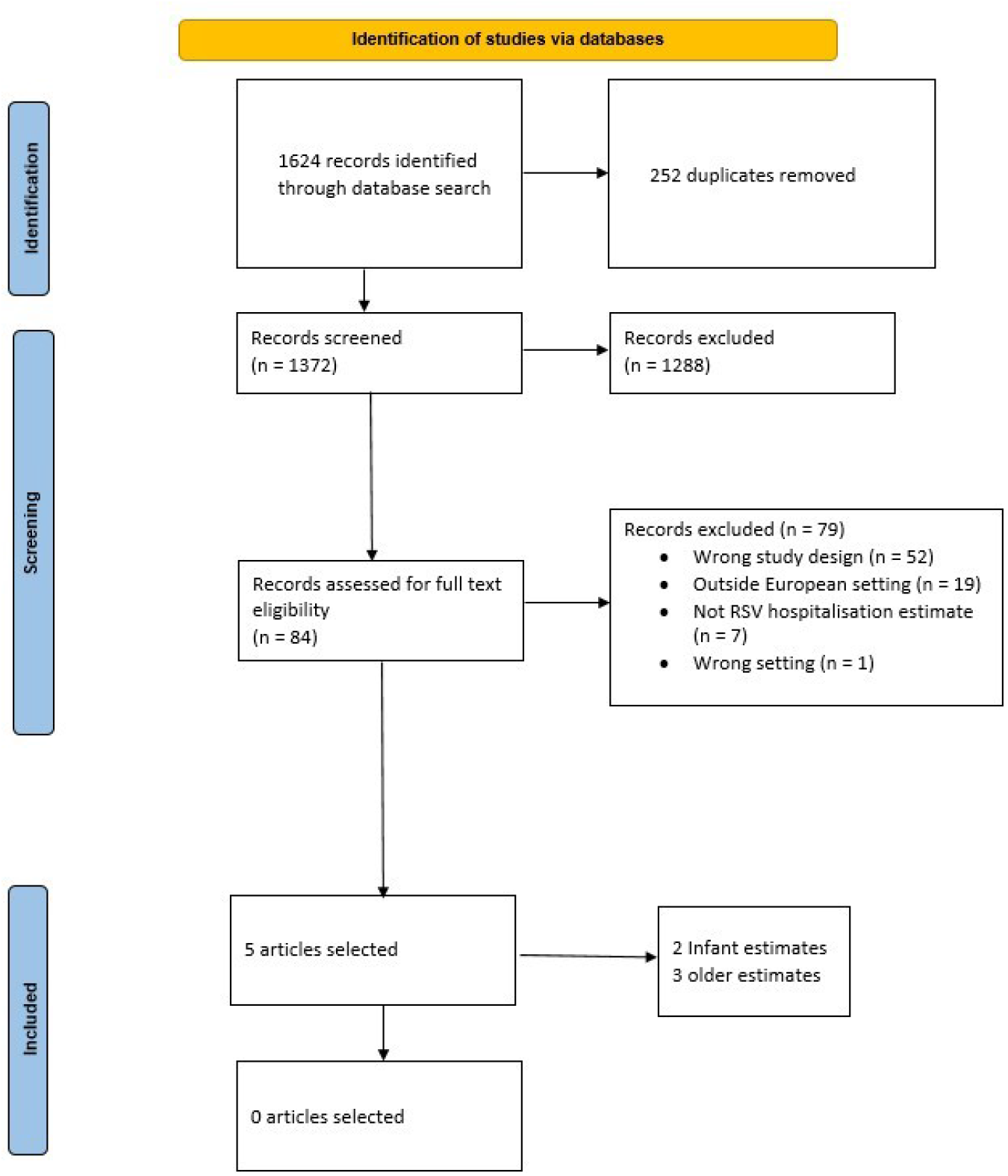
PRISMA flowchart outlining the search records of RSV-associated hospitalisation estimates in European countries.

**Figure 2:**
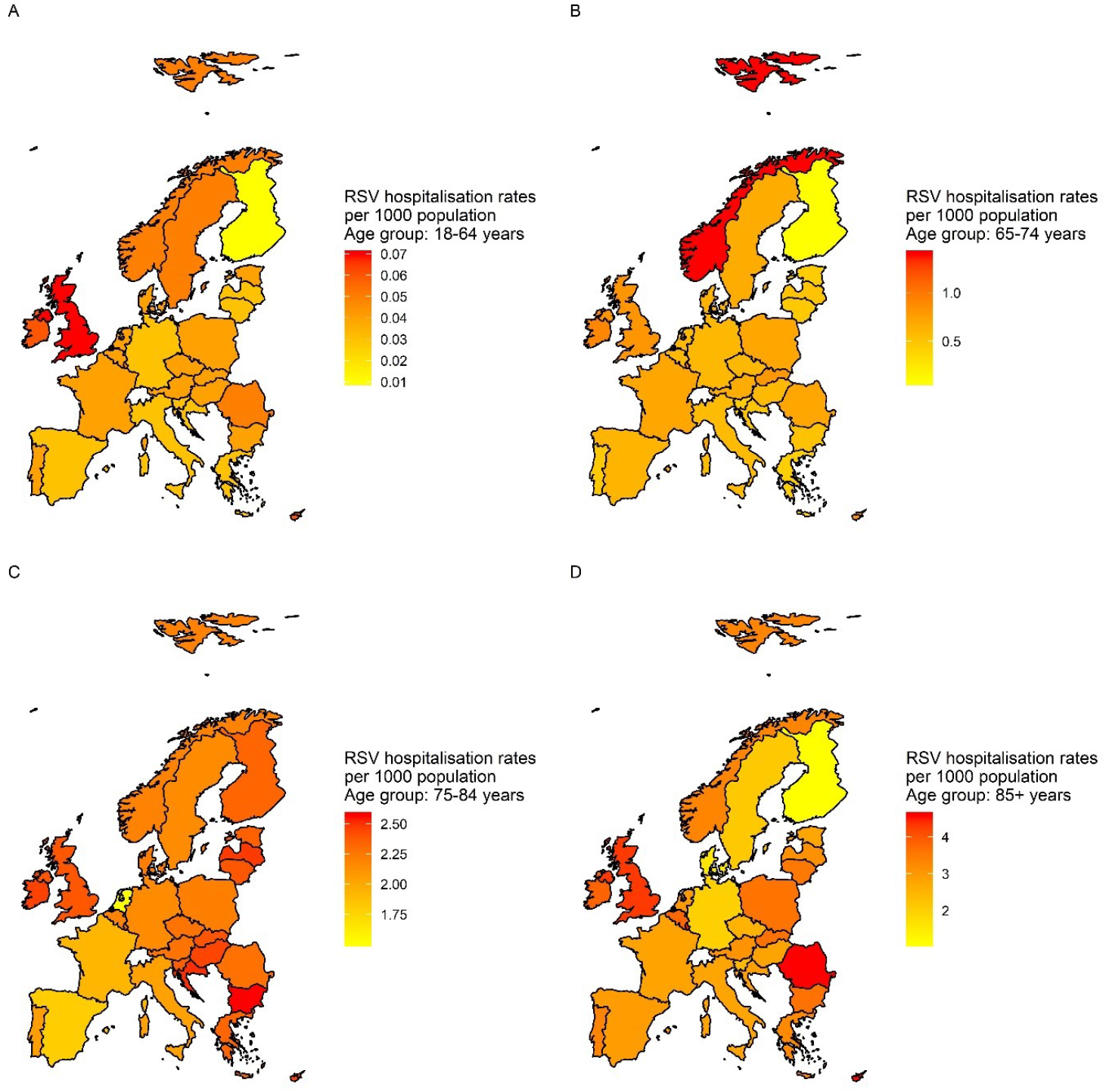
RSV-associated hospitalisation rates per 1000 population in 28 EU countries and Norway. A: RSV-associated hospitalisation rates per 1 000 in adults aged 18-64 years. B: RSV-associated hospitalisation rates per 1 000 in adults aged 65-74 years. C: RSV-associated hospitalisation rates per 1 000 in adults aged 75-84 years. D: RSV-associated hospitalisation rates per 1 000 in adults aged ≥85 years.

### Stage 2 estimates

#### Number of RSV-associated hospital admissions in 28 EU countries

Extrapolating the data from five EU countries and Norway to 28 EU countries (including the UK), we estimate that on average 158 229 (95% Confidence Interval (CI): 140 865 – 175 592) RSV-associated hospitalisations occurred annually among adults aged 18 years and older (Table 2). The highest average annual number of RSV-associated hospital admissions among adults aged 18-64 years: 2 896 (95% CI: 2 039 – 3 752), and ≥85 years: 6 002 (95% CI: 4 028 – 7 976) were estimated to occur in the UK whilst the highest average annual RSV-associated number of hospital admissions for adults aged 65-74 years: 5 387 (95% CI: 3 885 – 6 889) and 75-84 years: 13 843 (95% CI: 11 923 – 15 764) was estimated to occur in Germany (Table 2). The lowest average annual RSV-associated number of hospital admissions in all age groups occurs in Malta (Table 2). Of the overall RSV-associated hospital admissions, 145 102 (95%CI: 129 961-160 242) occurred among adults older than 65 years per year. Among adults aged 75-84 years, we estimate an average annual RSV-associated hospital admission of 74 519 (95% CI: 69 923 – 79 115) at a rate of 2.24 (95% CI: 2.10-2.38) per 1 000 adults per year (Table 3). The highest proportion of RSV-associated hospitalisations (47%) occurred in this age group in the EU as well as in all countries compared to other age groups (Table 4). In the 28 EU countries, 91.7% of all adult RSV-associated hospital admissions occurred in adults older than 65 years (Table 4).

**Table 2:** Estimated average annual RSV-associated hospitalisations by age groups in 28 EU countries (including the UK) and Norway

| Average annual number of hospitalisations (95% confidence interval) per age group |  |  |  |  |  |
| --- | --- | --- | --- | --- | --- |
| Country | 18-64 years | 65-74 years | 75-84 years | ≥85 years | ≥65 years |
| **EU 28\*** | 13127 (10904, 15350) | 32679 (27594, 37764) | 74519 (69923, 79115) | 37904 (32444, 43363) | 145102 (129961, 160242) |
| **Austria** | 202 (132, 272) | 587 (433, 740) | 1155 (1002, 1309) | 614 (434, 795) | 2356 (1869, 2844) |
| **Belgium** | 332 (244, 420) | 638 (456, 820) | 1608 (1396, 1820) | 1094 (852, 1335) | 3340 (2704, 3975) |
| **Bulgaria** | 207 (149, 265) | 494 (346, 641) | 1253 (1110, 1396) | 412 (308, 516) | 2159 (1764, 2553) |
| **Croatia** | 94 (60, 127) | 222 (146, 296) | 737 (648, 825) | 213 (151, 275) | 1172 (945, 1396) |
| **Cyprus** | 34 (27, 41) | 55 (42, 68) | 100 (88, 112) | 46 (36, 56) | 201 (166, 236) |
| **Czech Republic** | 305 (220, 391) | 736 (530, 941) | 1252 (1091, 1413) | 526 (374, 678) | 2514 (1995, 3032) |
| **Denmark** | 138 (94, 181) | 396 (284, 509) | 675 (584, 767) | 188 (89, 287) | 1259 (957, 1563) |
| **Estonia** | 35 (24, 45) | 62 (40, 85) | 207 (180, 234) | 84 (59, 108) | 353 (279, 427) |
| **Finland** | 26 (-16, 68) | 55 (-56, 166) | 795 (693, 896) | 150 (38, 262) | 1000 (675, 1324) |
| **France** | 1637 (1153, 2121) | 4244 (3172, 5316) | 7662 (6471, 8854) | 5901 (4258, 7543) | 17807 (13901, 21713) |
| **Germany** | 1393 (746, 2040) | 5387 (3885, 6889) | 13843 (11923, 15764) | 4514 (2713, 6314) | 23744 (18521, 28967) |
| **Greece** | 249 (164, 334) | 526 (331, 721) | 2133 (1876, 2390) | 967 (706, 1227) | 3626 (2913, 4338) |
| **Hungary** | 262 (181, 342) | 679 (498, 859) | 1332 (1163, 1500) | 512 (360, 664) | 2523 (2021, 3023) |
| **Ireland** | 214 (178, 251) | 322 (260, 385) | 459 (402, 514) | 224 (169, 279) | 1005 (831, 1178) |
| **Italy** | 1347 (876, 1817) | 3603 (2444, 4762) | 10531 (9126, 11936) | 4419 (2818, 6021) | 18553 (14388, 22719) |
| **Latvia** | 43 (27, 59) | 108 (72, 144) | 341 (299, 383) | 135 (100, 170) | 584 (471, 697) |
| **Lithuania** | 56 (33, 80) | 159 (109, 208) | 482 (421, 542) | 231 (177, 286) | 872 (707, 1036) |
| **Luxembourg** | 21 (16, 25) | 43 (35, 50) | 59 (51, 67) | 39 (30, 47) | 141 (116, 164) |
| **Malta** | 12 (9, 16) | 37 (29, 46) | 58 (50, 65) | 20 (13, 26) | 115 (92, 137) |
| **Netherlands** | 429 (296, 561) | 1000 (689, 1311) | 1427 (1150, 1704) | 1040 (751, 1328) | 3467 (2590, 4343) |
| **Norway** | 158 (117, 199) | 651 (558, 744) | 515 (441, 589) | 393 (295, 491) | 1559 (1294, 1824) |
| **Poland** | 970 (645, 1295) | 2254 (1685, 2823) | 4445 (3851, 5039) | 2246 (1690, 2802) | 8945 (7226, 10664) |
| **Portugal** | 252 (170, 333) | 543 (352, 734) | 1609 (1383, 1835) | 943 (712, 1173) | 3095 (2447, 3742) |
| **Romania** | 738 (576, 900) | 1422 (1100, 1745) | 2759 (2393, 3125) | 1430 (1162, 1699) | 5611 (4655, 6569) |
| **Slovakia** | 145 (98, 191) | 386 (305, 466) | 570 (501, 639) | 249 (189, 308) | 1205 (995, 1413) |
| **Slovenia** | 44 (27, 61) | 85 (51, 120) | 311 (272, 350) | 100 (64, 137) | 496 (387, 607) |
| **Spain** | 1031 (657, 1405) | 2509 (1742, 3275) | 5409 (4530, 6288) | 3217 (2133, 4301) | 11135 (8405, 13864) |
| **Sweden** | 285 (211, 360) | 681 (488, 876) | 1220 (1048, 1392) | 660 (445, 875) | 2561 (1981, 3143) |
| **United Kingdom** | 2896 (2039, 3752) | 4846 (2860, 6832) | 8625 (7034, 10216) | 6002 (4028, 7976) | 19473 (13922, 25024) |

\*Includes United Kingdom and excludes Norway

**Table 3:** Estimated rates of annual RSV-associated hospitalisations by age groups per 1 000 adults in 28 EU countries (including the UK) and Norway

| Country | Rate of hospitalisation per 1 000 adults (95% confidence interval) per age group |  |  |  |
| --- | --- | --- | --- | --- |
|  | 18-64 years | 65-74 years | 75-84 years | ≥85 years |
| **EU 28\*** | 0.04 (0.03, 0.05) | 0.66 (0.55, 0.76) | 2.24 (2.10, 2.38) | 2.99 (2.56, 3.42) |
| **Austria** | 0.04 (0.02, 0.05) | 0.69 (0.51, 0.87) | 2.23 (1.94, 2.53) | 2.86 (2.02, 3.70) |
| **Belgium** | 0.05 (0.04, 0.06) | 0.62 (0.45, 0.80) | 2.23 (1.94, 2.53) | 3.79 (2.95, 4.62) |
| **Bulgaria** | 0.05 (0.03, 0.06) | 0.59 (0.42, 0.77) | 2.59 (2.29, 2.88) | 3.30 (2.46, 4.14) |
| **Croatia** | 0.04 (0.02, 0.05) | 0.53 (0.35, 0.70) | 2.46 (2.16, 2.75) | 2.87 (2.03, 3.70) |
| **Cyprus** | 0.06 (0.05, 0.07) | 0.76 (0.58, 0.93) | 2.53 (2.23, 2.82) | 3.94 (3.10, 4.77) |
| **Czech Republic** | 0.04 (0.03, 0.06) | 0.64 (0.46, 0.82) | 2.30 (2.00, 2.59) | 2.89 (2.05, 3.72) |
| **Denmark** | 0.04 (0.03, 0.05) | 0.63 (0.45, 0.81) | 2.19 (1.89, 2.49) | 1.60 (0.76, 2.44) |
| **Estonia** | 0.04 (0.03, 0.05) | 0.49 (0.31, 0.67) | 2.28 (1.99, 2.58) | 2.87 (2.03, 3.71) |
| **Finland** | 0.01 (0.00, 0.02) | 0.09 (-.09, 0.27) | 2.32 (2.03, 2.62) | 1.12 (0.28, 1.96) |
| **France** | 0.04 (0.03, 0.06) | 0.70 (0.53, 0.88) | 1.90 (1.60, 2.19) | 3.01 (2.18, 3.85) |
| **Germany** | 0.03 (0.01, 0.04) | 0.64 (0.46, 0.82) | 2.13 (1.83, 2.42) | 2.10 (1.26, 2.94) |
| **Greece** | 0.04 (0.02, 0.05) | 0.48 (0.30, 0.66) | 2.45 (2.15, 2.74) | 3.10 (2.27, 3.94) |
| **Hungary** | 0.04 (0.03, 0.05) | 0.67 (0.49, 0.85) | 2.33 (2.04, 2.63) | 2.82 (1.99, 3.66) |
| **Ireland** | 0.07 (0.06, 0.09) | 0.92 (0.74, 1.10) | 2.42 (2.13, 2.72) | 3.37 (2.54, 4.21) |
| **Italy** | 0.04 (0.02, 0.05) | 0.55 (0.38, 0.73) | 2.21 (1.92, 2.51) | 2.31 (1.47, 3.14) |
| **Latvia** | 0.03 (0.02, 0.05) | 0.54 (0.36, 0.71) | 2.40 (2.11, 2.70) | 3.22 (2.38, 4.05) |
| **Lithuania** | 0.03 (0.02, 0.04) | 0.57 (0.39, 0.75) | 2.35 (2.05, 2.64) | 3.58 (2.75, 4.42) |
| **Luxembourg** | 0.06 (0.04, 0.07) | 1.02 (0.84, 1.20) | 2.14 (1.84, 2.44) | 3.82 (2.99, 4.66) |
| **Malta** | 0.04 (0.03, 0.06) | 0.78 (0.60, 0.96) | 2.35 (2.05, 2.65) | 2.64 (1.80, 3.49) |
| **Netherlands** | 0.04 (0.03, 0.05) | 0.58 (0.40, 0.76) | 1.53 (1.23, 1.83) | 3.02 (2.18, 3.86) |
| **Norway** | 0.05 (0.04, 0.06) | 1.37 (1.18, 1.57) | 2.10 (1.80, 2.41) | 3.42 (2.56, 4.27) |
| **Poland** | 0.04 (0.03, 0.05) | 0.70 (0.53, 0.88) | 2.21 (1.91, 2.51) | 3.38 (2.54, 4.21) |
| **Portugal** | 0.04 (0.03, 0.05) | 0.51 (0.33, 0.68) | 2.10 (1.80, 2.40) | 3.43 (2.59, 4.26) |
| **Romania** | 0.06 (0.05, 0.07) | 0.78 (0.61, 0.96) | 2.22 (1.93, 2.52) | 4.45 (3.61, 5.28) |
| **Slovakia** | 0.04 (0.03, 0.05) | 0.85 (0.68, 1.03) | 2.43 (2.13, 2.72) | 3.48 (2.65, 4.32) |
| **Slovenia** | 0.03 (0.02, 0.05) | 0.44 (0.26, 0.62) | 2.37 (2.07, 2.66) | 2.32 (1.48, 3.15) |
| **Spain** | 0.03 (0.02, 0.05) | 0.58 (0.40, 0.76) | 1.82 (1.52, 2.11) | 2.48 (1.65, 3.32) |
| **Sweden** | 0.05 (0.04, 0.06) | 0.63 (0.45, 0.81) | 2.11 (1.81, 2.41) | 2.58 (1.74, 3.42) |
| **United Kingdom** | 0.07 (0.05, 0.09) | 0.77 (0.46, 1.09) | 2.31 (1.89, 2.74) | 3.95 (2.65, 5.25) |

\*Includes United Kingdom and excludes Norway

**Table 4:** Proportion of overall RSV-associated hospitalisations occurring in adults aged 18-64 years, 65-74 years, 75-84 years and ≥85 years in the EU and in each country

| Country | 18-64 years | 65-74 years | 75-84 years | ≥85 years | ≥65 years |
| --- | --- | --- | --- | --- | --- |
| **EU 28\*** | 8.3 | 20.7 | 47.1 | 24.0 | 91.7 |
| **Austria** | 7.9 | 22.9 | 45.2 | 24.0 | 92.1 |
| **Belgium** | 9.0 | 17.4 | 43.8 | 29.8 | 91.0 |
| **Bulgaria** | 8.7 | 20.9 | 53.0 | 17.4 | 91.3 |
| **Croatia** | 7.4 | 17.5 | 58.2 | 16.8 | 92.6 |
| **Cyprus** | 14.5 | 23.4 | 42.6 | 19.6 | 85.5 |
| **Czech Republic** | 10.8 | 26.1 | 44.4 | 18.7 | 89.2 |
| **Denmark** | 9.9 | 28.3 | 48.3 | 13.5 | 90.1 |
| **Estonia** | 9.0 | 16.0 | 53.4 | 21.6 | 91.0 |
| **Finland** | 2.5 | 5.4 | 77.5 | 14.6 | 97.5 |
| **France** | 8.4 | 21.8 | 39.4 | 30.3 | 91.6 |
| **Germany** | 5.5 | 21.4 | 55.1 | 18.0 | 94.5 |
| **Greece** | 6.4 | 13.6 | 55.0 | 25.0 | 93.6 |
| **Hungary** | 9.4 | 24.4 | 47.8 | 18.4 | 90.6 |
| **Ireland** | 17.6 | 26.4 | 37.7 | 18.4 | 82.4 |
| **Italy** | 6.8 | 18.1 | 52.9 | 22.2 | 93.2 |
| **Latvia** | 6.9 | 17.2 | 54.4 | 21.5 | 93.1 |
| **Lithuania** | 6.0 | 17.1 | 51.9 | 24.9 | 94.0 |
| **Luxembourg** | 13.0 | 26.5 | 36.4 | 24.1 | 87.0 |
| **Malta** | 9.4 | 29.1 | 45.7 | 15.7 | 90.6 |
| **Netherlands** | 11.0 | 25.7 | 36.6 | 26.7 | 89.0 |
| **Norway** | 9.2 | 37.9 | 30.0 | 22.9 | 90.8 |
| **Poland** | 9.8 | 22.7 | 44.8 | 22.7 | 90.2 |
| **Portugal** | 7.5 | 16.2 | 48.1 | 28.2 | 92.5 |
| **Romania** | 11.6 | 22.4 | 43.5 | 22.5 | 88.4 |
| **Slovakia** | 10.7 | 28.6 | 42.2 | 18.4 | 89.3 |
| **Slovenia** | 8.1 | 15.7 | 57.6 | 18.5 | 91.9 |
| **Spain** | 8.5 | 20.6 | 44.5 | 26.4 | 91.5 |
| **Sweden** | 10.0 | 23.9 | 42.9 | 23.2 | 90.0 |
| **United Kingdom** | 12.9 | 21.7 | 38.6 | 26.8 | 87.1 |

\*Includes United Kingdom and excludes Norway

Importantly, the two model outputs did not differ significantly between the nearest-neighbour matching and multiple imputation approaches using the two indicator sets. The model only slightly differed by age group and the multiple imputation model estimates were slightly higher than the nearest-neighbour matching models.

#### Rate of RSV-associated hospital admissions per age group and across 28 EU countries

The average annual RSV-associated hospital admission rates and numbers varied across the EU. The estimated rate of hospitalisation was higher for adults ≥85 years (2.99, 95% CI: 2.56-3.42 per 1 000) with an average number of hospital admission of 37 904 (95%CI: 32 444 – 43 363) per year. Persons aged 18-64 years had the lowest hospitalisation rate of 0.04 (95% CI: 0.03-0.05 per 1 000 per year) and average annual hospitalisation of 13 127 (95% CI: 10 904 – 15 350).

The highest hospital admission rate per 1 000 adults aged 18-64 years [0.07 (95% CI: 0.06 −0.09)] was estimated in Ireland; the highest rates per 1 000 adults aged 65-74 years [1.37 (95% CI: 1.18 - 1.57)], 75-84 years [2.59 (95% CI: 2.29 - 2.88)] and ≥85 years [4.45 (95% CI: 3.61 - 5.28)] were estimated for Norway, Bulgaria and Romania respectively. The lowest hospital admission rates per 1 000 adults aged 18-64 years [0.01 (95% CI: -.00 - 0.02)], 65-74 years [0.09 (95% CI: -.09 - 0.27)] and ≥85 years [1.12 (95% CI: 0.28 - 1.96)] were estimated in Finland, whilst the lowest hospital admission rate in adults aged 75-84 years [1.53 (95% CI: 1.23 - 1.83) per 1 000] was estimated in the Netherlands (Table 3).

## Discussion

RSV infection among older adults, especially those with underlying health conditions, is an important cause of acute respiratory infections that can lead to hospitalisation. A previous multi-site RSV burden cohort study conducted among healthy older adults and adults with comorbidities in three European countries showed that RSV burden in both healthy older adults and adults with comorbidities is substantial and comparable to influenza [24].

These estimates of RSV-associated hospitalisations in older adults are the first analysis integrating available data in six European countries to provide empirical evidence of the disease burden in this population across the EU. Our estimates show that RSV causes a high annual number of hospital admissions in adults across the EU (roughly 160 000 per year) with about 92% of cases occurring in adults aged 65 years and older. The highest annual count of RSV-associated hospitalisations occurred among adults aged 75-84 years and the highest rate of RSV-associated hospital admissions occurred among adults over 85 years. It is important to note that for a condition which was considered in the past to be primarily a disease of young children, the average RSV-associated hospitalisation estimate in adults was lower, but of a similar magnitude to that in children aged 0-4 years: 158 229 (95%CI: 140 865 – 175 592) versus 245 244 (95%CI: 224 688 – 265 799) hospitalisations per year [25]. Indeed, taking the estimates for children aged 0-4 years into account; we estimate that 39% (145 604 out of 371 299) of the annual number of RSV-associated hospitalisations in the EU occurred in persons aged 65 years and older. With the changing demographics across Europe where elderly populations are increasing in size and considering the fact that RSV hospitalisation in adults can lead to acute functional decline and reduced quality of life [26], there is a need to get better data and estimates of the true burden of RSV in this age group.

Our estimates, based on the available data, suggest a high burden of RSV in terms of hospital admissions in adult populations. Several studies have previously reported the burden and severity of clinical outcomes of RSV among adults in different settings [7, 9, 10, 27-30]. For instance, in previous studies, the prevalence of RSV has been reported to be two times higher in patients aged over 75 years compared to those aged below 60 years [31]. Sundaram et al. report that RSV is a major cause of acute respiratory infection in adults aged over 50 years and RSV infection is more frequently associated with adults aged 65-79 years when compared to those aged 50-64 years [32]. In a community cohort of adults aged 50 years and over, the incidence of medically attended RSV infection was found to increase with age and the highest incidence occurred among persons aged over 70 years [33]. Our estimates of RSV-associated hospitalisation rates are consistent with the most recent pooled estimates based on prospective surveillance and modelling from the USA which report annual rates of RSV-associated hospitalisation of 178 (152-204) per 100 000 adults aged ≥65 years [34]. Among adults aged ≥65 years in the US, about 159 000 RSV-associated hospitalisations are estimated to occur each year which is comparable to our estimates of over 145 000 hospitalisations occurring in the same age group across the EU [34]. We have focused on RSV-associated hospital admission in older adults which is considered more severe, expensive and has quality of life implications [35], but recognise that there is also a significant amount of RSV disease burden in the outpatient setting [36].

Our study had several important limitations. The national estimates used for the extrapolations were generated mainly from Northern and Western European countries. Additional national estimates from Southern and Eastern Europe would yield more representative and reliable estimates for the EU. The two sets of ten indicators used to produce the extrapolations were selected based on their availability in all the countries included and are not (always) specific to RSV. Future estimates generated with this approach should attempt to use a more RSV-aligned set of indicators (e.g., indoor and outdoor pollution, geographical and ecological factors, etc.). The estimates used for Stage 1 were based on the overall number of hospitalisations with respiratory infections along with weekly test positives from laboratory data. The number of admissions and positive tests differed between the included countries, and as such, the estimates are still affected by country- or age-specific coding practices. Our estimates may differ from other studies using a similar approach because the Stage 1 results used depended on overall, non-age-specific virology data and the regression models were built with seasonal trends and polynomials [18]. It would be useful to provide estimates by year, as has been done for influenza-associated respiratory mortality for the EU [37], to assess temporal trends in RSV-associated hospitalisations and to produce more recent estimates to better understand how the COVID-19 pandemic has influenced RSV activity and its burdens such as infections, hospitalisations, and deaths.

There is also a lot of uncertainty due to low proportions of cases tested for RSV. These uncertainties may be mitigated only by using the observed number of admissions with a specific diagnosis or if laboratory confirmation of diagnosis (all cases tested), and lower sensitivity of testing methods in this age group are improved by combining polymerase chain reaction (PCR) and serology. With RSV, the reported burden of disease is broadly considered too low, a problem attributed to low levels of routine testing being used, and imperfect reporting of hospitalisation ICD-10 codes [38, 39]. In addition, routine diagnostic testing may not be appropriate for adults with RSV due to lower viral loads in adults compared to children [28, 40]. While the estimated burden of disease might be generally more uncertain, the high proportion of RSV-associated hospital admissions is more likely to reflect the overall healthcare burden and daily experiences of clinical professionals. The difference may even be larger in the elderly as this group is more often hospitalised due to other respiratory reasons than RSV and testing for RSV is probably done in only a limited number of elderly patients.

We have based this work on regression model estimates [which are not so dependent on high levels of RSV testing] to increase the comparability of the Stage 1 estimates [41] and to limit the risk of underestimating the RSV disease burden in adults (as there is less testing in this age group)[13]. Considering that the review of the literature found no eligible recent studies, there is a need for more data from more countries to generate improved estimates with less uncertainty as the uncertainty intervals given here are falsely narrow and do not capture all sources of uncertainty.

The main strength of this work is that we have used national data from several countries in a single agreed analysis framework to estimate the RSV-associated hospital admission burden for the EU. Our estimates provide a key insight into the healthcare burden that may be associated with RSV infection and can be used to inform healthcare planning, priority setting and resource allocation. Our results are important for public health policy and practice as these estimates may guide development of RSV surveillance systems, provide baseline evidence for the introduction of future vaccines, and raise awareness to generate more granular data. These data should therefore contribute to decision-making and policy formulation to improve relevant prevention, diagnostics and healthcare service delivery for this population.

## Data Availability

All data produced in the present study are available upon reasonable request to the authors and/or the RESCEU Steering Committee

## Footnote Page

### The RESCEU investigators are as follows

Harish NAIR (University of Edinburgh), Harry CAMPBELL (University of Edinburgh), Philippe Beutels (Universiteit Antwerpen), Louis Bont (University Medical Center Utrecht), Andrew Pollard (University of Oxford), Peter Openshaw (Imperial College London), Federico Martinon-Torres (Servicio Galego de Saude), Terho Heikkinen (University of Turku and Turku University Hospital), Adam Meijer (National Institute for Public Health and the Environment), Thea K. Fischer (Statens Serum Institut), Maarten van den Berge (University of Groningen), Carlo Giaquinto (PENTA Foundation), Michael Abram (AstraZeneca), Kena Swanson (Pfizer), Bishoy Rizkalla (GlaxoSmithKline), Charlotte Vernhes (Sanofi Pasteur), Scott Gallichan (Sanofi Pasteur), Jeroen Aerssens (Janssen), Veena Kumar (Novavax), Eva Molero (Team-It Research).

### Financial support

This work is part of RESCEU. RESCEU has received funding from the Innovative Medicines Initiative 2 Joint Undertaking under grant agreement No 116019. This Joint Undertaking receives support from the European Union’s Horizon 2020 research and innovation programme and EFPIA. This publication only reflects the author’s view and the JU is not responsible for any use that may be made of the information it contains herein.

### Disclaimer

Data from the Norwegian Patient Registry have been used in this publication. The interpretation and reporting of these data are the sole responsibility of the authors, and no endorsement by the Norwegian Patient Registry is intended nor should be inferred. This work reflects only the author’s views and opinions. The EC is not responsible for any use that may be made of the information it contains.

