## Supplementary data for "Estimation of the number of RSV-associated hospitalisations in adults in the European Union"

**Supplementary File 1:** Search strategy for the literature review

**Medline**

1. exp Respiratory Syncytial Virus Infections/ or exp Respiratory Syncytial Viruses/ or RSV.mp. or exp Respiratory Syncytial Virus, Human/

2. respiratory syncytial virus*.mp.

3. exp Pneumonia/ or exp Pneumonia, Viral/ or pneumonia.mp.

4. bronchiolitis.mp. or exp Bronchiolitis/ or exp Bronchiolitis, Viral/

5. exp Respiratory Tract Infections/ or respiratory infection*.mp.

6. respiratory disease*.mp. or exp Respiratory Tract Diseases/

7. exp Hospital Mortality/ or exp Mortality/ or mortality.mp.

8. exp Death/ or death*.mp. or exp "Cause of Death"/

9. exp Hospitalization/ or hospitalisation*.mp.

10. burden.mp.

11. exp Epidemiology/ or epidemiology.mp.

12. exp Models, Statistical/ or statistical model*.mp.

13. model*.mp.

14. estimat*.mp.

15. modelling estimate*.mp.

16. 1 or 2

17. 3 or 4 or 5 or 6

18. 7 or 8 or 9 or 10 or 11

19. 12 or 13 or 14 or 15

20. 16 and 17 and 18 and 19

21. limit 20 to (da="1995 - 20200127" and (female or humans or male))

**EMBASE**

1. exp Respiratory syncytial pneumovirus/ or RSV.mp. or exp respiratory syncytial virus infection/

2. respiratory syncytial virus*.mp.

3. exp pneumonia/ or exp community acquired pneumonia/ or exp infectious pneumonia/ or pneumonia.mp. or exp virus pneumonia/

4. bronchiolitis.mp. or exp bronchiolitis/ or exp viral bronchiolitis/

5. exp respiratory tract infection/ or respiratory infection*.mp.

6. exp respiratory tract disease/ or respiratory disease*.mp.

7. exp hospital mortality/ or mortality.mp. or exp mortality/ or exp mortality rate/

8. exp death/ or death*.mp. or exp "cause of death"/

9. exp hospitalization/ or hospitalisation*.mp.

10. burden.mp.

11. exp epidemiology/ or epidemiology.mp.

12. exp statistical model/ or statistical model*.mp. or exp model/

13. model*.mp.

14. estimat*.mp.

15. modelling estimate*.mp.

16. 1 or 2

17. 3 or 4 or 5 or 6

18. 7 or 8 or 9 or 10 or 11

19. 12 or 13 or 14 or 15

20. 16 and 17 and 18 and 19

21. limit 20 to (da="1995 - 20200127" and "humans only (removes records about animals)")

**Supplementary File 2:** Set of indicators used for the Stage 2 extrapolation

Indicator set 1:

-	% total population age 0-14

-	% total population age 60plus

-	death rate in specific age group (first three age groups death rate 0 years)

-	% primary and lower secondary education adult population

-	houshold income

-	% deprived adult population

-	% working in hightech jobs

-	% manual working

-	% unemployed age 20-64

-	agricultural output

Indicator set 2:

-	% total population age 0-14

-	% total population age 60plus

-	death rate in specific age group (first three age groups deathrate 0 years)

-	% primary and lower secondary education adult population

-	household income

-	curative hospital beds per 100000 population

-	% health and social workers of total workers

-	industrial crop

-	fruits production

-	youth unemployment

Sources:

-	Population and death-rates come from the Human Mortality database. Available at https://www.mortality.org/ [Last accessed 01 March 2022].

-	Eurostat. Available at: https://ec.europa.eu/eurostat/data/database [Last accessed 01 March 2022].

**Supplementary File 3:** RSV-associated hospitalisation rates per 1,000 population per age group per year calculated using the nearest neighbour matching approach and indicator set 1

| Country | 18-64 years | 65-74 years | 75-84 years | ≥85 years |
| --- | --- | --- | --- | --- |
| EU 28* | 0.03 (0.03, 0.04) | 0.53 (0.44, 0.62) | 2.30 (2.17, 2.43) | 2.80 (2.47, 3.13) |
| Austria | 0.04 (0.02, 0.05) | 0.61 (0.39, 0.82) | 2.20 (1.87, 2.53) | 2.48 (1.68, 3.28) |
| Belgium | 0.04 (0.02, 0.05) | 0.46 (0.25, 0.67) | 2.11 (1.78, 2.43) | 2.73 (1.94, 3.52) |
| Bulgaria | 0.02 (0.01, 0.04) | 0.46 (0.25, 0.67) | 2.43 (2.11, 2.76) | 2.76 (1.97, 3.55) |
| Croatia | 0.02 (0.01, 0.04) | 0.53 (0.32, 0.75) | 2.50 (2.17, 2.82) | 3.05 (2.26, 3.84) |
| Cyprus | 0.04 (0.03, 0.05) | 0.63 (0.42, 0.84) | 2.28 (1.95, 2.60) | 3.29 (2.50, 4.08) |
| Czech Republic | 0.03 (0.02, 0.04) | 0.49 (0.28, 0.70) | 2.59 (2.26, 2.91) | 3.07 (2.28, 3.86) |
| Denmark | 0.03 (0.02, 0.04) | 0.45 (0.23, 0.67) | 2.26 (1.93, 2.59) | 2.14 (1.34, 2.94) |
| Estonia | 0.03 (0.02, 0.05) | 0.51 (0.30, 0.72) | 2.49 (2.16, 2.81) | 2.57 (1.77, 3.38) |
| Finland | 0.01 (0.00, 0.03) | 0.14 (-0.08, 0.36) | 2.18 (1.85, 2.51) | 1.88 (1.08, 2.68) |
| France | 0.03 (0.02, 0.05) | 0.64 (0.43, 0.85) | 2.14 (1.82, 2.47) | 2.48 (1.68, 3.28) |
| Germany | 0.03 (0.01, 0.04) | 0.54 (0.33, 0.75) | 2.32 (1.99, 2.64) | 2.71 (1.90, 3.51) |
| Greece | 0.03 (0.01, 0.04) | 0.43 (0.22, 0.64) | 2.26 (1.93, 2.58) | 3.02 (2.23, 3.81) |
| Hungary | 0.03 (0.02, 0.04) | 0.55 (0.34, 0.77) | 2.50 (2.17, 2.82) | 3.01 (2.22, 3.80) |
| Ireland | 0.04 (0.03, 0.05) | 0.52 (0.31, 0.73) | 2.45 (2.13, 2.78) | 2.80 (2.01, 3.59) |
| Italy | 0.03 (0.01, 0.04) | 0.49 (0.28, 0.71) | 2.24 (1.91, 2.56) | 2.71 (1.92, 3.50) |
| Latvia | 0.02 (0.01, 0.04) | 0.42 (0.20, 0.63) | 2.50 (2.17, 2.82) | 2.92 (2.13, 3.71) |
| Lithuania | 0.02 (0.01, 0.04) | 0.38 (0.17, 0.59) | 2.51 (2.19, 2.84) | 2.73 (1.94, 3.52) |
| Luxembourg | 0.04 (0.03, 0.06) | 0.68 (0.46, 0.90) | 2.00 (1.67, 2.33) | 2.93 (2.13, 3.73) |
| Malta | 0.04 (0.02, 0.05) | 0.59 (0.38, 0.81) | 2.43 (2.10, 2.76) | 2.94 (2.14, 3.74) |
| Netherlands | 0.04 (0.03, 0.05) | 0.55 (0.33, 0.77) | 1.75 (1.42, 2.08) | 2.92 (2.12, 3.72) |
| Norway | 0.05 (0.03, 0.06) | 1.22 (0.99, 1.45) | 2.24 (1.90, 2.58) | 3.32 (2.50, 4.13) |
| Poland | 0.03 (0.01, 0.04) | 0.53 (0.32, 0.74) | 2.41 (2.09, 2.74) | 3.01 (2.22, 3.80) |
| Portugal | 0.02 (0.01, 0.04) | 0.35 (0.14, 0.56) | 2.32 (2.00, 2.65) | 2.83 (2.04, 3.62) |
| Romania | 0.03 (0.01, 0.04) | 0.51 (0.30, 0.72) | 2.24 (1.91, 2.56) | 3.07 (2.28, 3.86) |
| Slovakia | 0.03 (0.01, 0.04) | 0.56 (0.35, 0.78) | 2.40 (2.08, 2.73) | 3.02 (2.23, 3.81) |
| Slovenia | 0.02 (0.01, 0.04) | 0.42 (0.21, 0.63) | 2.47 (2.15, 2.80) | 2.57 (1.78, 3.36) |
| Spain | 0.03 (0.02, 0.05) | 0.53 (0.31, 0.74) | 2.06 (1.73, 2.38) | 2.76 (1.97, 3.55) |
| Sweden | 0.05 (0.03, 0.06) | 0.76 (0.54, 0.98) | 2.18 (1.85, 2.51) | 2.61 (1.81, 3.41) |
| United Kingdom | 0.08 (0.06, 0.10) | 0.71 (0.43, 0.99) | 2.60 (2.22, 2.99) | 4.76 (3.90, 5.63) |

*Includes United Kingdom and excludes Norway

**Supplementary File 4:** RSV-associated hospitalisation rates per 1,000 population per age group per year calculated using the nearest neighbour matching approach and indicator set 2

| Country | 18-64 years | 65-74 years | 75-84 years | ≥85 years |
| --- | --- | --- | --- | --- |
| EU 28* | 0.04 (0.03, 0.04) | 0.65 (0.57, 0.73) | 2.03 (1.91, 2.14) | 2.82 (2.52, 3.13) |
| Austria | 0.04 (0.03, 0.05) | 0.71 (0.53, 0.89) | 1.93 (1.62, 2.23) | 2.97 (2.18, 3.76) |
| Belgium | 0.04 (0.03, 0.05) | 0.63 (0.45, 0.81) | 1.84 (1.53, 2.14) | 3.04 (2.26, 3.83) |
| Bulgaria | 0.03 (0.02, 0.05) | 0.63 (0.45, 0.82) | 2.18 (1.88, 2.49) | 3.17 (2.38, 3.95) |
| Croatia | 0.03 (0.02, 0.05) | 0.50 (0.32, 0.69) | 2.17 (1.86, 2.47) | 3.04 (2.25, 3.82) |
| Cyprus | 0.04 (0.03, 0.05) | 0.57 (0.38, 0.75) | 2.05 (1.75, 2.36) | 3.20 (2.41, 3.99) |
| Czech Republic | 0.04 (0.02, 0.05) | 0.65 (0.46, 0.83) | 2.18 (1.88, 2.49) | 3.05 (2.26, 3.84) |
| Denmark | 0.05 (0.03, 0.06) | 0.75 (0.56, 0.93) | 1.90 (1.60, 2.21) | 2.00 (1.21, 2.79) |
| Estonia | 0.03 (0.02, 0.05) | 0.50 (0.32, 0.68) | 2.26 (1.95, 2.56) | 2.44 (1.66, 3.23) |
| Finland | 0.01 (0.00, 0.02) | 0.17 (-.01, 0.35) | 2.00 (1.70, 2.31) | 1.67 (0.88, 2.45) |
| France | 0.04 (0.03, 0.05) | 0.69 (0.50, 0.87) | 1.91 (1.61, 2.22) | 2.88 (2.09, 3.66) |
| Germany | 0.03 (0.02, 0.05) | 0.62 (0.44, 0.80) | 1.93 (1.62, 2.23) | 2.75 (1.96, 3.53) |
| Greece | 0.03 (0.02, 0.05) | 0.56 (0.38, 0.74) | 1.99 (1.69, 2.29) | 3.04 (2.26, 3.83) |
| Hungary | 0.04 (0.03, 0.05) | 0.66 (0.48, 0.84) | 2.18 (1.88, 2.49) | 3.19 (2.41, 3.98) |
| Ireland | 0.05 (0.04, 0.06) | 0.79 (0.61, 0.97) | 2.22 (1.91, 2.52) | 2.95 (2.17, 3.74) |
| Italy | 0.04 (0.03, 0.05) | 0.68 (0.49, 0.86) | 1.91 (1.61, 2.22) | 2.53 (1.75, 3.32) |
| Latvia | 0.03 (0.02, 0.05) | 0.64 (0.45, 0.82) | 2.26 (1.95, 2.56) | 3.03 (2.25, 3.82) |
| Lithuania | 0.04 (0.02, 0.05) | 0.65 (0.46, 0.83) | 2.18 (1.88, 2.49) | 2.80 (2.02, 3.59) |
| Luxembourg | 0.05 (0.03, 0.06) | 0.80 (0.61, 0.98) | 1.90 (1.60, 2.21) | 2.91 (2.14, 3.69) |
| Malta | 0.02 (0.01, 0.04) | 0.72 (0.53, 0.90) | 2.13 (1.82, 2.45) | 2.61 (1.81, 3.40) |
| Netherlands | 0.04 (0.03, 0.05) | 0.61 (0.43, 0.80) | 1.58 (1.28, 1.89) | 2.98 (2.19, 3.76) |
| Norway | 0.05 (0.04, 0.06) | 1.20 (0.97, 1.44) | 1.89 (1.57, 2.21) | 2.89 (2.06, 3.71) |
| Poland | 0.04 (0.02, 0.05) | 0.64 (0.45, 0.82) | 2.08 (1.77, 2.38) | 3.05 (2.26, 3.83) |
| Portugal | 0.02 (0.01, 0.03) | 0.54 (0.36, 0.73) | 2.00 (1.70, 2.31) | 2.83 (2.04, 3.62) |
| Romania | 0.04 (0.03, 0.05) | 0.61 (0.43, 0.80) | 1.91 (1.61, 2.22) | 3.11 (2.32, 3.89) |
| Slovakia | 0.04 (0.03, 0.05) | 0.70 (0.52, 0.88) | 2.16 (1.85, 2.46) | 3.11 (2.33, 3.90) |
| Slovenia | 0.03 (0.02, 0.04) | 0.44 (0.26, 0.62) | 2.24 (1.94, 2.55) | 2.64 (1.86, 3.43) |
| Spain | 0.04 (0.03, 0.06) | 0.70 (0.52, 0.88) | 1.82 (1.52, 2.13) | 2.63 (1.85, 3.42) |
| Sweden | 0.05 (0.04, 0.06) | 0.84 (0.66, 1.03) | 1.93 (1.62, 2.23) | 2.54 (1.76, 3.33) |
| United Kingdom | 0.08 (0.06, 0.10) | 0.70 (0.45, 0.96) | 2.32 (1.96, 2.68) | 4.27 (3.35, 5.19) |

*Includes United Kingdom and excludes Norway

**Supplementary File 5:** RSV-associated hospitalisation rates per 1,000 population per age group per year calculated using the multiple imputation approach and indicator set 1

| Country | 18-64 years | 65-74 years | 75-84 years | ≥85 years |
| --- | --- | --- | --- | --- |
| EU 28* | 0.04 (0.04, 0.05) | 0.69 (0.59, 0.80) | 2.31 (2.16, 2.45) | 3.05 (2.46, 3.65) |
| Austria | 0.03 (0.02, 0.04) | 0.77 (0.63, 0.91) | 2.58 (2.30, 2.87) | 2.71 (1.85, 3.58) |
| Belgium | 0.06 (0.05, 0.07) | 0.78 (0.64, 0.92) | 2.43 (2.15, 2.71) | 4.11 (3.25, 4.98) |
| Bulgaria | 0.08 (0.07, 0.09) | 0.78 (0.63, 0.92) | 2.90 (2.61, 3.18) | 3.53 (2.67, 4.39) |
| Croatia | 0.03 (0.02, 0.05) | 0.52 (0.38, 0.66) | 2.29 (2.01, 2.58) | 2.19 (1.33, 3.06) |
| Cyprus | 0.08 (0.06, 0.09) | 0.75 (0.61, 0.89) | 2.83 (2.55, 3.12) | 4.05 (3.19, 4.91) |
| Czech Republic | 0.06 (0.05, 0.07) | 0.84 (0.70, 0.99) | 2.29 (2.01, 2.57) | 2.87 (2.01, 3.74) |
| Denmark | 0.04 (0.03, 0.05) | 0.57 (0.42, 0.71) | 2.40 (2.11, 2.68) | 1.05 (0.18, 1.91) |
| Estonia | 0.03 (0.02, 0.05) | 0.30 (0.16, 0.44) | 2.27 (1.98, 2.55) | 3.29 (2.42, 4.15) |
| Finland | 0.00 (-.01, 0.01) | 0.03 (-.11, 0.17) | 2.38 (2.10, 2.67) | 0.36 (-.50, 1.22) |
| France | 0.03 (0.02, 0.05) | 0.73 (0.59, 0.87) | 1.59 (1.30, 1.87) | 3.22 (2.36, 4.09) |
| Germany | 0.02 (0.01, 0.03) | 0.67 (0.53, 0.81) | 2.17 (1.88, 2.45) | 1.18 (0.32, 2.04) |
| Greece | 0.03 (0.02, 0.05) | 0.59 (0.45, 0.73) | 2.59 (2.30, 2.87) | 3.59 (2.73, 4.45) |
| Hungary | 0.05 (0.04, 0.06) | 0.65 (0.51, 0.79) | 2.47 (2.19, 2.76) | 2.57 (1.71, 3.43) |
| Ireland | 0.08 (0.07, 0.10) | 0.90 (0.76, 1.04) | 2.31 (2.02, 2.59) | 1.35 (0.49, 2.21) |
| Italy | 0.03 (0.01, 0.04) | 0.69 (0.54, 0.83) | 1.77 (1.49, 2.05) | 1.47 (0.61, 2.33) |
| Latvia | 0.03 (0.02, 0.04) | 0.39 (0.25, 0.53) | 2.60 (2.32, 2.89) | 4.51 (3.65, 5.37) |
| Lithuania | 0.02 (0.01, 0.04) | 0.69 (0.54, 0.83) | 2.58 (2.30, 2.87) | 5.23 (4.37, 6.09) |
| Luxembourg | 0.05 (0.04, 0.06) | 0.98 (0.84, 1.12) | 2.52 (2.24, 2.81) | 4.55 (3.69, 5.41) |
| Malta | 0.05 (0.04, 0.06) | 0.67 (0.52, 0.81) | 2.62 (2.34, 2.90) | 1.60 (0.74, 2.46) |
| Netherlands | 0.04 (0.03, 0.05) | 0.58 (0.44, 0.72) | 1.41 (1.13, 1.69) | 3.08 (2.22, 3.94) |
| Norway | 0.05 (0.04, 0.06) | 1.53 (1.39, 1.68) | 1.95 (1.67, 2.23) | 4.32 (3.45, 5.18) |
| Poland | 0.04 (0.03, 0.05) | 0.81 (0.67, 0.95) | 2.14 (1.85, 2.42) | 4.84 (3.98, 5.71) |
| Portugal | 0.07 (0.05, 0.08) | 0.58 (0.44, 0.73) | 2.26 (1.98, 2.54) | 4.91 (4.05, 5.77) |
| Romania | 0.10 (0.09, 0.11) | 1.00 (0.86, 1.14) | 2.37 (2.09, 2.65) | 6.94 (6.07, 7.80) |
| Slovakia | 0.05 (0.03, 0.06) | 1.22 (1.08, 1.36) | 2.31 (2.03, 2.59) | 3.18 (2.32, 4.04) |
| Slovenia | 0.03 (0.02, 0.04) | 0.47 (0.33, 0.61) | 2.48 (2.20, 2.77) | 1.20 (0.34, 2.06) |
| Spain | 0.02 (0.00, 0.03) | 0.53 (0.39, 0.67) | 1.68 (1.39, 1.96) | 0.87 (0.01, 1.73) |
| Sweden | 0.05 (0.03, 0.06) | 0.37 (0.22, 0.51) | 2.34 (2.06, 2.63) | 2.68 (1.82, 3.54) |
| United Kingdom | 0.06 (0.04, 0.09) | 0.79 (0.47, 1.11) | 2.09 (1.62, 2.55) | 2.89 (1.05, 4.73) |

*Includes United Kingdom and excludes Norway

**Supplementary File 6:** RSV-associated hospitalisation rates per 1,000 population per age group per year calculated using the multiple imputation approach and indicator set 2

| Country | 18-64 years | 65-74 years | 75-84 years | ≥85 years |
| --- | --- | --- | --- | --- |
| EU 28* | 0.05 (0.04, 0.06) | 0.75 (0.62, 0.88) | 2.32 (2.17, 2.48) | 3.29 (2.80, 3.79) |
| Austria | 0.03 (0.02, 0.05) | 0.67 (0.50, 0.85) | 2.22 (1.95, 2.49) | 3.29 (2.38, 4.20) |
| Belgium | 0.06 (0.04, 0.07) | 0.62 (0.44, 0.79) | 2.56 (2.29, 2.83) | 5.26 (4.35, 6.17) |
| Bulgaria | 0.04 (0.03, 0.06) | 0.51 (0.33, 0.68) | 2.84 (2.57, 3.11) | 3.75 (2.84, 4.65) |
| Croatia | 0.05 (0.04, 0.06) | 0.54 (0.37, 0.72) | 2.86 (2.59, 3.13) | 3.19 (2.28, 4.10) |
| Cyprus | 0.09 (0.08, 0.10) | 1.08 (0.90, 1.25) | 2.94 (2.67, 3.20) | 5.21 (4.31, 6.12) |
| Czech Republic | 0.05 (0.04, 0.07) | 0.57 (0.39, 0.75) | 2.13 (1.86, 2.40) | 2.55 (1.65, 3.46) |
| Denmark | 0.04 (0.03, 0.06) | 0.77 (0.60, 0.95) | 2.20 (1.93, 2.47) | 1.21 (0.30, 2.12) |
| Estonia | 0.07 (0.06, 0.08) | 0.66 (0.48, 0.83) | 2.12 (1.85, 2.39) | 3.19 (2.28, 4.10) |
| Finland | 0.00 (-.01, 0.02) | 0.02 (-.16, 0.20) | 2.73 (2.46, 3.00) | 0.56 (-.34, 1.47) |
| France | 0.06 (0.05, 0.08) | 0.76 (0.58, 0.93) | 1.95 (1.68, 2.22) | 3.47 (2.57, 4.38) |
| Germany | 0.03 (0.01, 0.04) | 0.73 (0.55, 0.90) | 2.10 (1.83, 2.37) | 1.79 (0.88, 2.69) |
| Greece | 0.05 (0.04, 0.07) | 0.34 (0.17, 0.52) | 2.96 (2.69, 3.23) | 2.77 (1.86, 3.68) |
| Hungary | 0.04 (0.03, 0.06) | 0.82 (0.64, 0.99) | 2.18 (1.91, 2.44) | 2.51 (1.61, 3.42) |
| Ireland | 0.12 (0.11, 0.13) | 1.47 (1.29, 1.64) | 2.70 (2.44, 2.97) | 6.38 (5.47, 7.29) |
| Italy | 0.05 (0.04, 0.06) | 0.36 (0.18, 0.53) | 2.93 (2.66, 3.20) | 2.52 (1.61, 3.43) |
| Latvia | 0.05 (0.04, 0.06) | 0.70 (0.53, 0.88) | 2.26 (1.99, 2.53) | 2.41 (1.50, 3.32) |
| Lithuania | 0.04 (0.02, 0.05) | 0.58 (0.41, 0.76) | 2.11 (1.84, 2.38) | 3.56 (2.66, 4.47) |
| Luxembourg | 0.08 (0.07, 0.09) | 1.62 (1.44, 1.80) | 2.13 (1.86, 2.39) | 4.90 (3.99, 5.81) |
| Malta | 0.06 (0.05, 0.07) | 1.14 (0.97, 1.32) | 2.20 (1.94, 2.47) | 3.43 (2.52, 4.33) |
| Netherlands | 0.04 (0.03, 0.05) | 0.57 (0.40, 0.75) | 1.37 (1.10, 1.64) | 3.12 (2.21, 4.03) |
| Norway | 0.05 (0.04, 0.06) | 1.53 (1.35, 1.71) | 2.34 (2.07, 2.61) | 3.15 (2.24, 4.06) |
| Poland | 0.04 (0.03, 0.06) | 0.85 (0.67, 1.02) | 2.21 (1.95, 2.48) | 2.61 (1.70, 3.52) |
| Portugal | 0.05 (0.04, 0.06) | 0.55 (0.38, 0.73) | 1.81 (1.54, 2.08) | 3.13 (2.22, 4.04) |
| Romania | 0.07 (0.05, 0.08) | 1.01 (0.84, 1.19) | 2.37 (2.10, 2.64) | 4.68 (3.77, 5.58) |
| Slovakia | 0.05 (0.03, 0.06) | 0.93 (0.76, 1.11) | 2.84 (2.57, 3.10) | 4.62 (3.71, 5.53) |
| Slovenia | 0.05 (0.04, 0.06) | 0.42 (0.24, 0.60) | 2.28 (2.01, 2.55) | 2.85 (1.94, 3.76) |
| Spain | 0.05 (0.03, 0.06) | 0.57 (0.39, 0.74) | 1.71 (1.44, 1.97) | 3.66 (2.76, 4.57) |
| Sweden | 0.05 (0.04, 0.07) | 0.56 (0.38, 0.73) | 1.99 (1.72, 2.26) | 2.47 (1.56, 3.38) |
| United Kingdom | 0.07 (0.04, 0.09) | 0.89 (0.48, 1.29) | 2.25 (1.75, 2.75) | 3.88 (2.31, 5.45) |

*Includes United Kingdom and excludes Norway

**Supplementary File 7:** RSV-associated average annual hospitalisations per age group per year calculated using the nearest neighbour matching approach and indicator set 1

| Country | 18-64 years | 65-74 years | 75-84 years | ≥85 years |
| --- | --- | --- | --- | --- |
| EU 28* | 9857 (7924, 11790) | 26607 (22149, 31065) | 76606 (72273, 80938) | 35437 (31264, 39610) |
| Austria | 210 (134, 286) | 516 (329, 703) | 1138 (966, 1310) | 532 (360, 704) |
| Belgium | 246 (151, 341) | 473 (256, 689) | 1516 (1282, 1750) | 788 (560, 1016) |
| Bulgaria | 100 (37, 163) | 381 (205, 557) | 1177 (1020, 1335) | 344 (246, 442) |
| Croatia | 65 (29, 102) | 225 (135, 314) | 750 (652, 847) | 226 (167, 284) |
| Cyprus | 22 (14, 30) | 46 (30, 61) | 90 (77, 103) | 38 (29, 48) |
| Czech Republic | 209 (116, 302) | 565 (321, 810) | 1410 (1232, 1587) | 560 (416, 703) |
| Denmark | 108 (60, 155) | 283 (146, 420) | 696 (594, 799) | 252 (157, 346) |
| Estonia | 26 (15, 37) | 65 (38, 91) | 226 (196, 255) | 75 (51, 98) |
| Finland | 46 (1, 91) | 86 (-49, 221) | 745 (632, 859) | 252 (145, 359) |
| France | 1245 (721, 1768) | 3867 (2590, 5145) | 8654 (7342, 9967) | 4860 (3292, 6428) |
| Germany | 1419 (719, 2119) | 4539 (2750, 6329) | 15081 (12965, 17198) | 5804 (4085, 7523) |
| Greece | 169 (77, 261) | 469 (237, 701) | 1966 (1682, 2249) | 939 (693, 1185) |
| Hungary | 196 (109, 283) | 561 (346, 776) | 1427 (1241, 1612) | 546 (403, 690) |
| Ireland | 117 (78, 157) | 182 (108, 257) | 465 (403, 526) | 186 (134, 238) |
| Italy | 1032 (523, 1541) | 3210 (1830, 4591) | 10654 (9105, 12202) | 5182 (3671, 6694) |
| Latvia | 28 (11, 46) | 84 (41, 127) | 354 (308, 400) | 122 (89, 155) |
| Lithuania | 45 (19, 70) | 105 (46, 164) | 515 (449, 582) | 176 (125, 227) |
| Luxembourg | 16 (11, 21) | 28 (19, 38) | 55 (46, 64) | 30 (22, 38) |
| Malta | 10 (6, 14) | 28 (18, 39) | 60 (51, 68) | 22 (16, 27) |
| Netherlands | 417 (273, 560) | 948 (569, 1328) | 1633 (1323, 1944) | 1003 (727, 1278) |
| Norway | 148 (104, 193) | 579 (471, 688) | 548 (464, 631) | 381 (288, 475) |
| Poland | 714 (362, 1066) | 1696 (1017, 2374) | 4849 (4195, 5504) | 2001 (1476, 2526) |
| Portugal | 139 (50, 227) | 373 (146, 600) | 1779 (1530, 2028) | 779 (562, 996) |
| Romania | 361 (186, 537) | 928 (544, 1312) | 2776 (2373, 3180) | 986 (733, 1240) |
| Slovakia | 104 (54, 154) | 255 (159, 351) | 565 (488, 641) | 216 (159, 272) |
| Slovenia | 32 (14, 51) | 81 (40, 123) | 324 (281, 367) | 111 (77, 145) |
| Spain | 943 (538, 1348) | 2268 (1355, 3182) | 6127 (5158, 7095) | 3578 (2555, 4601) |
| Sweden | 274 (194, 355) | 819 (583, 1056) | 1260 (1068, 1453) | 669 (464, 874) |
| United Kingdom | 3218 (2540, 3895) | 4450 (2669, 6232) | 9704 (8267, 11140) | 7237 (5920, 8554) |

*Includes United Kingdom and excludes Norway

**Supplementary File 8:** RSV-associated average annual hospitalisations per age group per year calculated using the nearest neighbour matching approach and indicator set 2

| Country | 18-64 years | 65-74 years | 75-84 years | ≥85 years |
| --- | --- | --- | --- | --- |
| EU 28* | 11739 (9978, 13500) | 32367 (28171, 36562) | 67447 (63558, 71336) | 35771 (31888, 39653) |
| Austria | 235 (168, 301) | 605 (449, 760) | 997 (839, 1154) | 637 (468, 806) |
| Belgium | 279 (195, 363) | 646 (459, 833) | 1322 (1103, 1541) | 879 (651, 1106) |
| Bulgaria | 154 (98, 209) | 527 (375, 678) | 1056 (909, 1204) | 395 (297, 493) |
| Croatia | 90 (57, 122) | 212 (135, 289) | 651 (559, 742) | 225 (167, 284) |
| Cyprus | 23 (16, 29) | 41 (28, 54) | 81 (69, 93) | 37 (28, 47) |
| Czech Republic | 240 (158, 322) | 745 (534, 955) | 1189 (1023, 1355) | 555 (412, 699) |
| Denmark | 156 (114, 198) | 466 (352, 580) | 586 (492, 680) | 235 (142, 327) |
| Estonia | 28 (18, 38) | 63 (40, 86) | 205 (177, 233) | 71 (48, 94) |
| Finland | 36 (-4, 76) | 104 (-8, 216) | 685 (581, 789) | 224 (118, 329) |
| France | 1534 (1071, 1998) | 4139 (3040, 5238) | 7725 (6495, 8955) | 5633 (4094, 7172) |
| Germany | 1687 (1068, 2306) | 5219 (3679, 6758) | 12535 (10553, 14517) | 5892 (4205, 7579) |
| Greece | 233 (152, 314) | 616 (416, 816) | 1733 (1467, 1998) | 947 (703, 1192) |
| Hungary | 242 (165, 319) | 668 (483, 853) | 1245 (1071, 1419) | 579 (436, 722) |
| Ireland | 144 (109, 180) | 277 (213, 341) | 420 (362, 478) | 196 (144, 248) |
| Italy | 1559 (1109, 2009) | 4405 (3217, 5593) | 9110 (7660, 10560) | 4854 (3348, 6360) |
| Latvia | 42 (27, 58) | 128 (91, 165) | 320 (277, 363) | 127 (94, 160) |
| Lithuania | 65 (43, 88) | 179 (128, 230) | 448 (385, 510) | 181 (130, 232) |
| Luxembourg | 17 (12, 21) | 34 (26, 41) | 52 (44, 61) | 29 (22, 37) |
| Malta | 7 (4, 11) | 34 (25, 43) | 52 (45, 60) | 19 (13, 25) |
| Netherlands | 435 (309, 562) | 1060 (745, 1376) | 1477 (1193, 1761) | 1023 (753, 1294) |
| Norway | 157 (117, 198) | 571 (459, 682) | 463 (384, 541) | 332 (237, 427) |
| Poland | 952 (641, 1264) | 2032 (1448, 2615) | 4182 (3569, 4795) | 2026 (1503, 2549) |
| Portugal | 120 (42, 199) | 581 (386, 777) | 1534 (1301, 1767) | 779 (562, 995) |
| Romania | 481 (326, 636) | 1112 (781, 1443) | 2374 (1996, 2752) | 999 (746, 1252) |
| Slovakia | 143 (98, 187) | 315 (233, 397) | 506 (435, 578) | 222 (166, 278) |
| Slovenia | 41 (24, 57) | 86 (51, 122) | 294 (254, 334) | 114 (80, 149) |
| Spain | 1326 (968, 1684) | 3022 (2236, 3808) | 5434 (4527, 6342) | 3414 (2394, 4433) |
| Sweden | 284 (213, 355) | 910 (713, 1107) | 1114 (938, 1290) | 652 (450, 853) |
| United Kingdom | 3187 (2466, 3908) | 4412 (2798, 6026) | 8637 (7300, 9974) | 6490 (5089, 7891) |

*Includes United Kingdom and excludes Norway

**Supplementary File 9:** RSV-associated average annual hospitalisations per age group per year calculated using the multiple imputation approach and indicator set 1

| Country | 18-64 years | 65-74 years | 75-84 years | ≥85 years |
| --- | --- | --- | --- | --- |
| EU 28* | 14196 (11589, 16804) | 34477 (29306, 39648) | 76712 (71789, 81635) | 38666 (31151, 46181) |
| Austria | 179 (112, 245) | 653 (533, 774) | 1337 (1191, 1483) | 582 (397, 767) |
| Belgium | 415 (331, 498) | 800 (655, 945) | 1750 (1547, 1953) | 1188 (939, 1437) |
| Bulgaria | 377 (322, 432) | 645 (527, 762) | 1402 (1265, 1539) | 440 (333, 548) |
| Croatia | 89 (57, 121) | 220 (160, 279) | 688 (604, 773) | 163 (99, 227) |
| Cyprus | 42 (36, 49) | 54 (44, 64) | 112 (101, 123) | 47 (37, 57) |
| Czech Republic | 402 (321, 484) | 974 (811, 1138) | 1248 (1094, 1402) | 523 (366, 680) |
| Denmark | 136 (94, 177) | 354 (266, 443) | 739 (652, 826) | 123 (22, 224) |
| Estonia | 27 (17, 37) | 38 (20, 56) | 206 (180, 231) | 95 (70, 120) |
| Finland | 9 (-31, 48) | 17 (-70, 105) | 815 (718, 911) | 48 (-67, 164) |
| France | 1321 (862, 1781) | 4413 (3560, 5267) | 6404 (5264, 7544) | 6311 (4625, 7998) |
| Germany | 1160 (546, 1774) | 5640 (4445, 6836) | 14094 (12257, 15932) | 2526 (677, 4375) |
| Greece | 228 (147, 308) | 643 (488, 798) | 2253 (2007, 2499) | 1118 (850, 1387) |
| Hungary | 334 (257, 410) | 657 (513, 801) | 1412 (1251, 1574) | 467 (310, 623) |
| Ireland | 244 (210, 279) | 316 (266, 366) | 437 (383, 490) | 90 (33, 147) |
| Italy | 989 (543, 1435) | 4465 (3542, 5388) | 8422 (7077, 9766) | 2814 (1163, 4464) |
| Latvia | 36 (21, 51) | 79 (50, 107) | 369 (329, 409) | 188 (152, 224) |
| Lithuania | 45 (23, 67) | 190 (151, 229) | 530 (472, 588) | 338 (282, 394) |
| Luxembourg | 19 (14, 23) | 41 (35, 47) | 69 (62, 77) | 46 (37, 55) |
| Malta | 14 (10, 17) | 32 (25, 39) | 64 (57, 71) | 12 (5, 18) |
| Netherlands | 426 (300, 552) | 1001 (755, 1246) | 1316 (1053, 1580) | 1059 (762, 1355) |
| Norway | 161 (123, 200) | 728 (661, 795) | 477 (408, 546) | 496 (397, 595) |
| Poland | 1075 (766, 1384) | 2585 (2131, 3038) | 4296 (3728, 4864) | 3221 (2648, 3794) |
| Portugal | 424 (347, 502) | 626 (474, 778) | 1732 (1516, 1949) | 1351 (1114, 1588) |
| Romania | 1247 (1093, 1401) | 1813 (1556, 2070) | 2942 (2591, 3292) | 2231 (1954, 2508) |
| Slovakia | 165 (121, 209) | 551 (487, 615) | 543 (477, 609) | 227 (165, 288) |
| Slovenia | 38 (22, 54) | 92 (64, 120) | 325 (288, 363) | 52 (15, 89) |
| Spain | 491 (136, 846) | 2293 (1682, 2903) | 4994 (4153, 5835) | 1124 (7, 2241) |
| Sweden | 267 (197, 338) | 394 (241, 547) | 1355 (1191, 1518) | 686 (465, 907) |
| United Kingdom | 2470 (1457, 3483) | 4960 (2946, 6973) | 7775 (6031, 9520) | 4388 (1590, 7187) |

*Includes United Kingdom and excludes Norway

**Supplementary File 10:** RSV-associated average annual hospitalisations per age group per year calculated using the multiple imputation approach and indicator set 2

| Country | 18-64 years | 65-74 years | 75-84 years | ≥85 years |
| --- | --- | --- | --- | --- |
| EU 28* | 16715 (14125, 19306) | 37266 (30749, 43782) | 77310 (72070, 82550) | 41741 (35473, 48009) |
| Austria | 184 (113, 255) | 572 (422, 721) | 1149 (1010, 1288) | 706 (511, 901) |
| Belgium | 388 (299, 477) | 633 (453, 813) | 1844 (1651, 2037) | 1519 (1257, 1781) |
| Bulgaria | 197 (139, 256) | 421 (275, 567) | 1377 (1247, 1506) | 467 (354, 580) |
| Croatia | 130 (96, 164) | 229 (155, 303) | 859 (778, 939) | 236 (169, 304) |
| Cyprus | 50 (43, 57) | 78 (65, 91) | 116 (106, 127) | 61 (50, 72) |
| Czech Republic | 370 (283, 457) | 658 (455, 860) | 1162 (1016, 1308) | 465 (300, 631) |
| Denmark | 150 (106, 194) | 482 (372, 592) | 679 (596, 761) | 142 (35, 249) |
| Estonia | 57 (46, 67) | 83 (61, 106) | 192 (168, 217) | 93 (66, 119) |
| Finland | 13 (-29, 56) | 12 (-96, 120) | 933 (842, 1025) | 75 (-46, 197) |
| France | 2446 (1957, 2936) | 4557 (3499, 5615) | 7865 (6783, 8948) | 6798 (5022, 8574) |
| Germany | 1306 (652, 1959) | 6148 (4666, 7631) | 13662 (11917, 15408) | 3833 (1886, 5780) |
| Greece | 366 (281, 452) | 375 (182, 567) | 2579 (2346, 2813) | 862 (579, 1144) |
| Hungary | 275 (193, 356) | 828 (650, 1007) | 1242 (1089, 1395) | 456 (291, 620) |
| Ireland | 351 (314, 388) | 514 (452, 575) | 512 (461, 563) | 424 (363, 484) |
| Italy | 1806 (1330, 2281) | 2331 (1187, 3475) | 13937 (12660, 15214) | 4827 (3089, 6564) |
| Latvia | 64 (48, 80) | 141 (106, 177) | 320 (282, 358) | 101 (63, 139) |
| Lithuania | 69 (45, 93) | 161 (112, 210) | 433 (378, 488) | 230 (172, 289) |
| Luxembourg | 30 (26, 35) | 68 (61, 75) | 58 (51, 66) | 50 (40, 59) |
| Malta | 18 (14, 21) | 55 (46, 63) | 54 (47, 61) | 25 (18, 32) |
| Netherlands | 436 (302, 570) | 989 (685, 1293) | 1281 (1030, 1531) | 1073 (761, 1385) |
| Norway | 164 (123, 205) | 726 (642, 809) | 573 (508, 639) | 362 (258, 466) |
| Poland | 1138 (809, 1467) | 2704 (2142, 3266) | 4452 (3913, 4992) | 1735 (1132, 2338) |
| Portugal | 323 (240, 405) | 591 (402, 779) | 1389 (1183, 1594) | 861 (611, 1111) |
| Romania | 862 (699, 1026) | 1835 (1517, 2154) | 2944 (2611, 3277) | 1504 (1213, 1796) |
| Slovakia | 166 (119, 213) | 421 (342, 500) | 666 (603, 729) | 330 (265, 394) |
| Slovenia | 66 (49, 83) | 82 (48, 116) | 299 (264, 334) | 124 (84, 163) |
| Spain | 1363 (985, 1741) | 2451 (1694, 3208) | 5080 (4281, 5879) | 4750 (3574, 5927) |
| Sweden | 315 (240, 390) | 602 (413, 792) | 1150 (995, 1305) | 633 (401, 866) |
| United Kingdom | 2708 (1693, 3722) | 5561 (3025, 8096) | 8385 (6538, 10231) | 5893 (3513, 8273) |

*Includes United Kingdom and excludes Norway
